# Performance Characteristics of the Abbott Architect SARS-CoV-2 IgG Assay and Seroprevalence Testing in Idaho

**DOI:** 10.1101/2020.04.27.20082362

**Authors:** Andrew Bryan, Gregory Pepper, Mark H. Wener, Susan L. Fink, Chihiro Morishima, Anu Chaudhary, Keith R. Jerome, Patrick C. Mathias, Alexander L. Greninger

**Author notes:** These authors contributed equally (Andrew Bryan, Gregory Pepper).

## Abstract

**Background:** Coronavirus disease-19 (COVID19), the novel respiratory illness caused by severe acute respiratory syndrome coronavirus 2 (SARS-CoV-2), is associated with severe morbidity and mortality. The rollout of diagnostic testing in the United States was slow, leading to numerous cases that were not tested for SARS-CoV-2 in February and March 2020, necessitating the use of serological testing to determine past infections.

**Methods:** We evaluated the Abbott SARS-CoV-2 IgG test for detection of anti-SARS-CoV-2 IgG antibodies by testing 3 distinct patient populations.

**Results:** We tested 1,020 serum specimens collected prior to SARS-CoV-2 circulation in the United States and found one false positive, indicating a specificity of 99.90%. We tested 125 patients who tested RT-PCR positive for SARS-CoV-2 for which 689 excess serum specimens were available and found sensitivity reached 100% at day 17 after symptom onset and day 13 after PCR positivity. Alternative index value thresholds for positivity resulted in 100% sensitivity and 100% specificity in this cohort. We tested 4,856 individuals from Boise, Idaho collected over one week in April 2020 as part of the Crush the Curve initiative and detected 87 positives for a positivity rate of 1.79%.

**Conclusions:** These data demonstrate excellent analytical performance of the Abbott SARS-CoV-2 IgG test as well as the limited circulation of the virus in the western United States. We expect the availability of high-quality serological testing will be a key tool in the fight against SARS-CoV-2.

## Introduction

Coronavirus disease-19 (COVID19) is a novel respiratory illness caused by Severe acute respiratory syndrome coronavirus 2 (SARS-CoV-2), a novel *Sarbecovirus* that recently emerged from Wuhan, China in late 2019 (1). COVID-19 often progresses to lower respiratory tract illness and can be associated with severe morbidity and mortality (2).

Serological testing can detect past cases of SARS-CoV-2 for which RT-PCR testing was either not performed or for which nasopharyngeal swab sampling resulted in false negatives. Serological tests require exceptional sensitivity and specificity, especially when seroprevalence is low, in order to have adequate positive predictive value (3). To date, most SARS-CoV-2 serological tests on the market have inadequate performance characteristics to perform widespread population or clinical testing (4). Here, we evaluated the Abbott SARS-CoV-2 IgG test for use on the Abbott Architect platform. This assay detects IgG antibodies against the SARS-CoV-2 nucleocapsid protein.

## Methods

### Patient Cohorts

Specificity samples were derived from de-identified excess serum specimens sent to our clinical virology laboratory in 2018 and 2019. Sensitivity specimens were derived from excess serum specimens sent for clinical testing from persons who tested RT-PCR positive for SARS-CoV-2 during March and April 2020. With the exception of the studies of biologic precision, for patients with an IgG result on more than 1 aliquot on a specific date following onset of symptoms or PCR positivity, only the mean index value for that patient-day was included in the data set to minimize the bias from individual patient seroconversion and variable numbers of samples per patient. For the calculations of sensitivity and specificity at the patient level using the manufacturer’s recommended index value cutoff of 1.40 (Figure 1), patients were assumed to be seronegative for each day preceding the most recent negative IgG result and to be seropositive for each day following an initial positive result. Serum specimens sent from the Boise, Idaho metropolitan area were collected over a one-week period in late April 2020 as part of the Crush the Curve initiative. This work was approved under a consent waiver by the University of Washington IRB.

**Figure 1.**
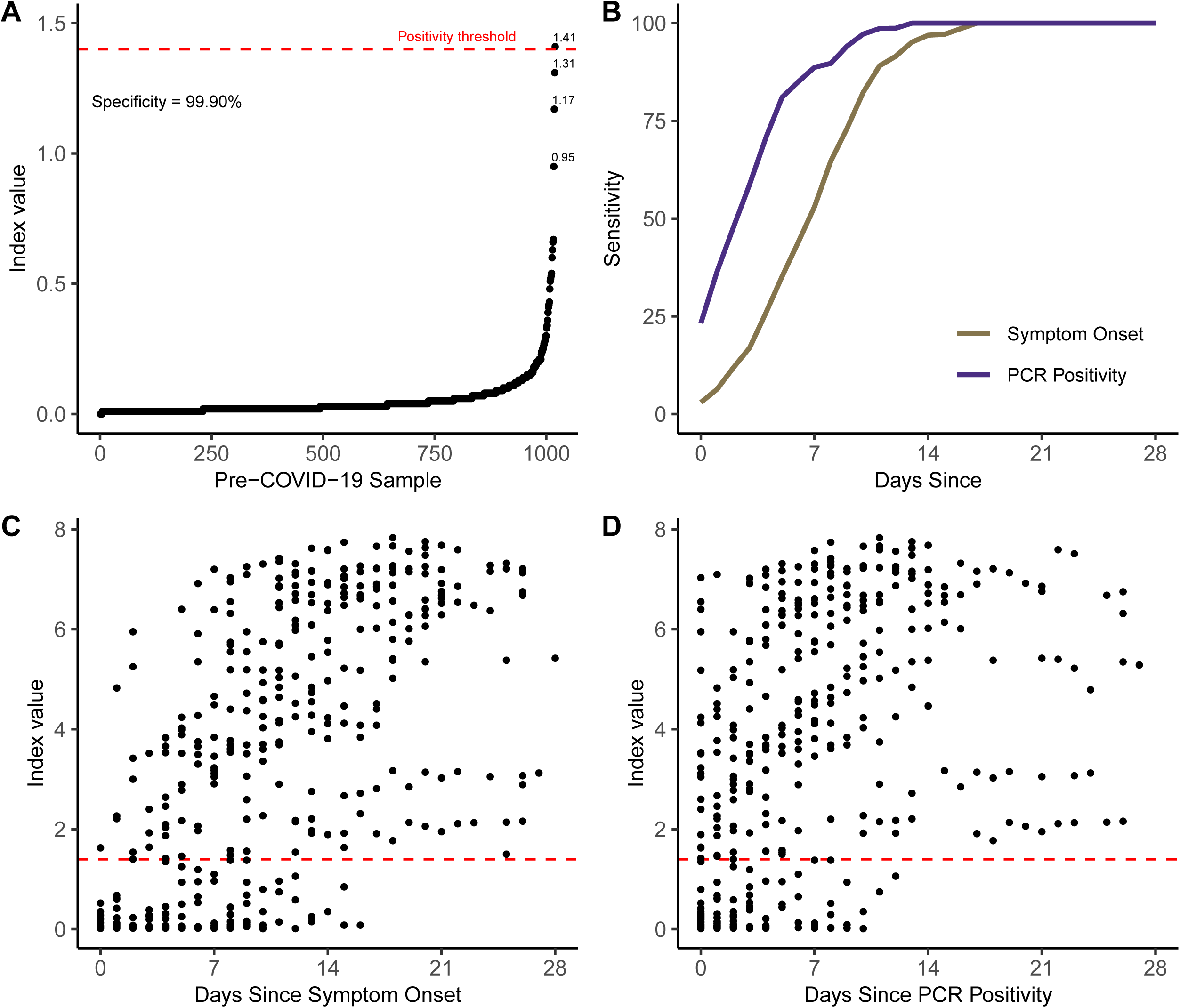
Performance characteristics of the Abbott SARS-CoV-2 IgG test. A) Specificity was determined using 1,020 serum specimens taken before circulation of SARS-CoV-2 in the United States. Index values by sample are shown in rank order, and sample with index values greater than 0.7 are labeled. B) Sensitivity by day since symptom onset and PCR positivity is depicted for 689 excess serum specimens comprising 415 unique patient follow-up days from 125 unique patients, using the manufacturer’s recommended positivity index value cutoff of 1.40. Index values are depicted by day since symptom onset (C) or PCR positivity (D). Index values were averaged for patients with multiple specimens from the same day. The index value threshold of 1.40 for positivity is depicted in the red horizontal dashed line.

### IgG Testing

Serum samples were run on the Abbott Architect instrument using the Abbott SARS-CoV-2 IgG assay after FDA notification following manufacturer’s instructions. The assay is a chemiluminescent microparticle immunoassay for qualitative detection of IgG in human serum or plasma against the SARS-CoV-2 nucleoprotein. The Architect requires a minimum of 100μL of serum or plasma. Qualitative results and index values reported by the instrument were used in analysis.

### Data analysis and visualization

Patient demographic information (sex and age) was extracted alongside laboratory order and result data (including index value) from the laboratory information system (Sunquest Laboratory, Tuscon, AZ). Partial AUC analysis and data visualization were performed using the R packages pROC, ggplot2, and cowplot (5, 6).

## Results

### Sensitivity and Specificity of the Abbott SARS-CoV-2 IgG Assay

To determine assay specificity, we used 1,020 deidentified serum specimens from 1,010 different individuals sent to our laboratory for HSV Western blot serology in 2018 and 2019, before SARS-CoV-2 was thought to be circulating in Washington State and the United States (7). One serum specimen from this set tested positive with an initial index value of 1.41 and repeated at 1.49, above the Abbott-determined positivity cutoff of 1.40 (Figure 1A). All other specimens tested negative, leading to an assay specificity of 99.90% in pre-COVID-19 serum.

To determine assay sensitivity, we used a series of 125 patients who tested RT-PCR positive for SARS-CoV-2 for which 689 excess serum specimens comprising 415 unique patient follow-up days were available. The vast majority of these patients were hospitalized at the University of Washington Medical Center-Northwest Campus in Seattle, WA between March and April 2020. Fifty-eight percent of patients were male and 42% female. The age distribution by decade of life was: 20–29: 2.4%, 30–39: 4.8%, 40–49: 9.6%, 50–59: 17.6%, 60–69: 17.6%, 70–79: 24.0%, 80–89: 16.0%, >= 90: 8.0%.

The sensitivity of the assay from the estimated day of symptom onset for the 125 patients included in our chart-review study was 53.1% (95%CI 39.4%-66.3%) at 7 days, 82.4% (51.0-76.4%) at 10 days, 96.9% (89.5-99.5%) at 14 days, and 100% (95.1%-100%) at day 17 using the manufacturer’s recommended cutoff of 1.4. The sensitivity from the date of PCR positivity was: 88.7% (78.5-94.4%) at 7 days, 97.2% (90.4-99.5%) at 10 days, 100.0% at 14 days (95.4-100.0%), and 100.0% (95.5-100.0%) at 17 days using the manufacturer’s recommended cutoff of 1.4. Intriguingly, 22 of 88 individuals (25%) for which serum was available on the first day of PCR positivity had simultaneous detection of serum anti-SARS-CoV-2 IgG and nasopharyngeal SARS-CoV-2 RNA (Figure 1B).

We next used our SARS-CoV-2 IgG index values over 415 unique patient-days to assess the change in index value over time, from the date of symptom onset (Figure 1C) and first positive PCR result (Figure 1D). For these patients early in the course of their infections, index values consistently increased over time, both on the review of individual patients with multiple IgG results over time and aggregate summary data.

Based on our data suggesting consistent seroconversion and the low false positive rate in our specificity study, we next asked what optimal index value cutoffs were for different days after onset of symptoms or PCR positivity. Partial AUC analysis was performed setting the minimum specificity between 99.0% and 99.9% (Tables 1–2), to minimize false positives given the low seroprevalence to SARS-CoV-2 expected in our population and to identify the optimal index thresholds for different potential uses of the test. These analyses indicated that optimal thresholds for the serologic diagnosis of SARS-CoV-2 was 1.42-1.49 at ≥ 17 days from symptom onset (sensitivity and specificity 100%); 0.7 at ≥ 14 days from onset (Sens 97.9%, Spec 99.6%); 0.7 at ≥ 10 days from onset (Sens 94.4%, Spec 99.6%); and 0.7 at ≥ 7 days from onset (Sens 88.0%, Spec 99.6%) (Figure 2).

**Table 1.** ROC analysis to determine optimal index value thresholds from day of onset.

| Minimum Specificity | Days From Onset | Threshold | Sensitivity | Specificity | pAUC |
| --- | --- | --- | --- | --- | --- |
| 99.9% | ≥ 17 | 1.5 | 100.0% | 100.0% | 100.0% |
|  | ≥ 14 | 1.5 | 97.2% | 100.0% | 98.6% |
|  | ≥ 10 | 1.5 | 92.1% | 100.0% | 96.1% |
|  | ≥ 7 | 1.4 | 84.2% | 100.0% | 92.1% |
| 99.8% | ≥ 17 | 1.5 | 100.0% | 100.0% | 100.0% |
|  | ≥ 14 | 1.5 | 97.2% | 100.0% | 98.6% |
|  | ≥ 10 | 1.5 | 92.1% | 100.0% | 96.1% |
|  | ≥ 7 | 1.3 | 94.9% | 99.9% | 92.2% |
| 99.5% | ≥ 17 | 1.5 | 100.0% | 100.0% | 100.0% |
|  | ≥ 14 | 0.8 | 97.9% | 99.6% | 98.6% |
|  | ≥ 10 | 0.7 | 94.4% | 99.6% | 96.4% |
|  | ≥ 7 | 0.7 | 88.0% | 99.6% | 92.8% |
| 99.0% | ≥ 17 | 1.5 | 100.0% | 100.0% | 100.0% |
|  | ≥ 14 | 0.8 | 97.9% | 99.6% | 98.8% |
|  | ≥ 10 | 0.6 | 94.9% | 99.3% | 96.9% |
|  | ≥ 7 | 0.6 | 88.4% | 99.3% | 93.4% |

**Table 2.** ROC analysis to determine optimal index value thresholds from day of first positive PCR result.

| Minimum Specificity | Days From PCR | Threshold | Sensitivity | Specificity | pAUC |
| --- | --- | --- | --- | --- | --- |
| 99.9% | ≥ 17 | 1.6 | 100.0% | 100.0% | 100.0% |
|  | ≥ 14 | 1.6 | 100.0% | 100.0% | 100.0% |
|  | ≥ 10 | 1.6 | 96.4% | 100.0% | 98.2% |
|  | ≥ 7 | 1.6 | 90.7% | 100.0% | 95.3% |
| 99.8% | ≥ 17 | 1.6 | 100.0% | 100.0% | 100.0% |
|  | ≥ 14 | 1.6 | 100.0% | 100.0% | 100.0% |
|  | ≥ 10 | 1.6 | 96.4% | 100.0% | 98.2% |
|  | ≥ 7 | 1.3 | 91.9% | 99.9% | 95.6% |
| 99.5% | ≥ 17 | 1.5 | 100.0% | 100.0% | 100.0% |
|  | ≥ 14 | 0.8 | 97.9% | 99.6% | 98.6% |
|  | ≥ 10 | 0.7 | 94.4% | 99.6% | 96.4% |
|  | ≥ 7 | 0.7 | 88.0% | 99.6% | 92.8% |
| 99.0% | ≥ 17 | 1.6 | 100.0% | 100.0% | 100.0% |
|  | ≥ 14 | 1.6 | 100.0% | 100.0% | 100.0% |
|  | ≥ 10 | 0.7 | 98.2% | 99.6% | 98.8% |
|  | ≥ 7 | 0.7 | 93.6% | 99.6% | 96.4% |

**Figure 2.**
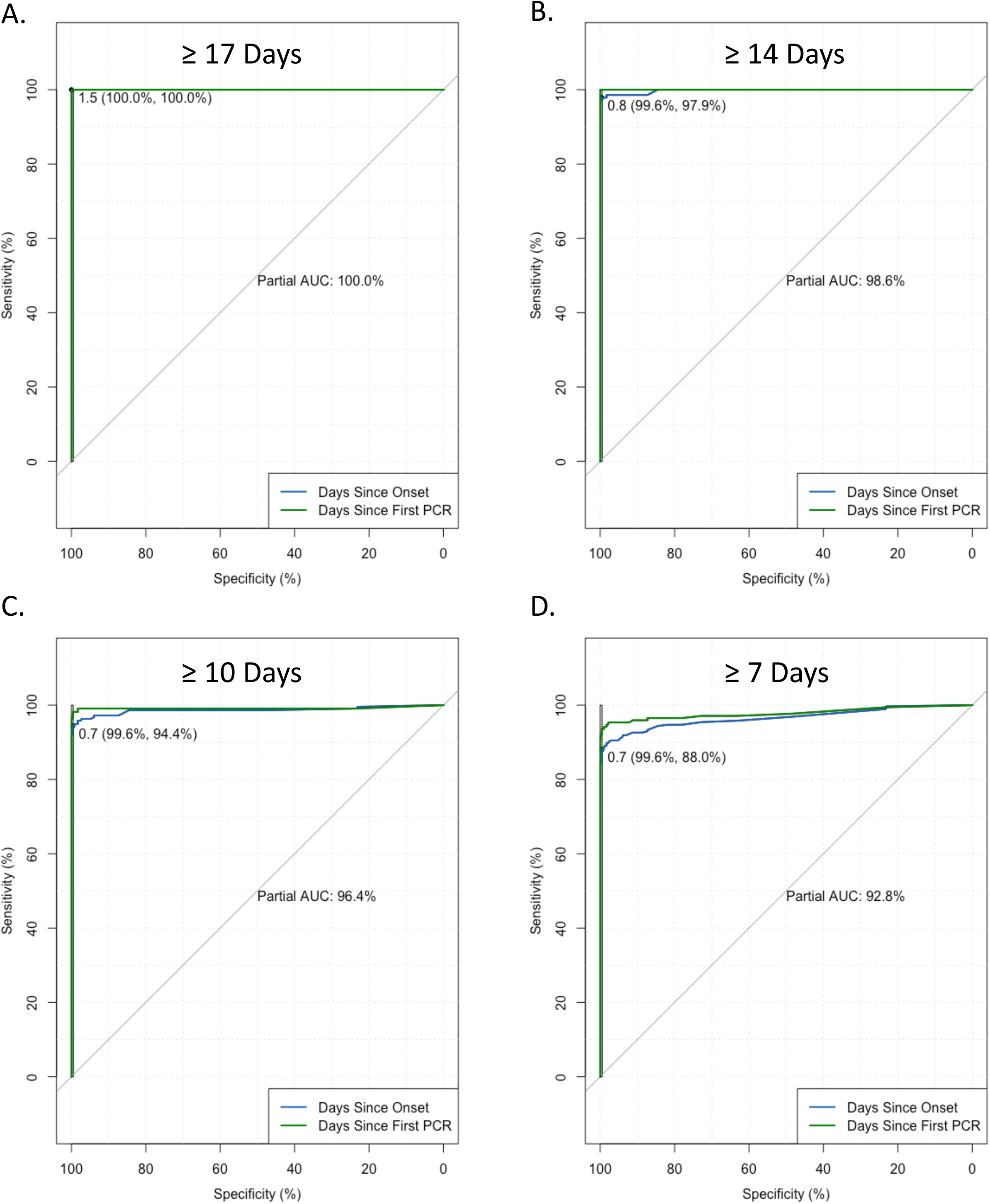
Receiver operating characteristic curves for the Abbott SARS-CoV-2 IgG test based on ≥ 17 days (A), ≥ 14 days (B), ≥ 10 days (C), ≥ 7 days (D) after symptom onset or PCR positivity. Minimum specificity was set to 99.5%.

Given our large unique data set, we next assessed the biologic variation of the antibody results in PCR positive patients by examining all test results where at least 3 remnant serum or plasma samples were available from the same day for the same patient. The coefficient of variation was calculated for each of 75 available patient-days and plotted against the index value (Figure 3A). The reproducibility of the measurable anti-SARS-CoV-2 IgG response was robust across the index value range except for 2 situations: 1) higher CV’s associated with very low index values <0.1 related to analytical ‘noise’; and 2) higher CVs related to the rapid change in antibody levels associated with active seroconversion. The CV was < 10% for all included patient-days above an index value of 0.4 except for 4 data points representing 3 different patients in the process of seroconversion. For 3 of these 4 patient-days with CVs >10%, samples had been drawn several hours apart. To further examine the process of seroconversion in individual patients, we identified 7 patients that had IgG results available on at least 5 different patient-days and in whom we captured the process of seroconversion, plus 1 patient that appeared to be in the process of seroconverting, but did not cross positivity threshold (Figure 3B). In addition to assessment of the biologic variation, traditional analytic precision was determined: the same remnant sample was analyzed 5-10 times on each of 3 Abbott Architect instruments, yielding individuals CVs of between 1.4% and 2.5% and a cumulative CV of 2.6% (cumulative mean 2.26).

**Figure 3.**
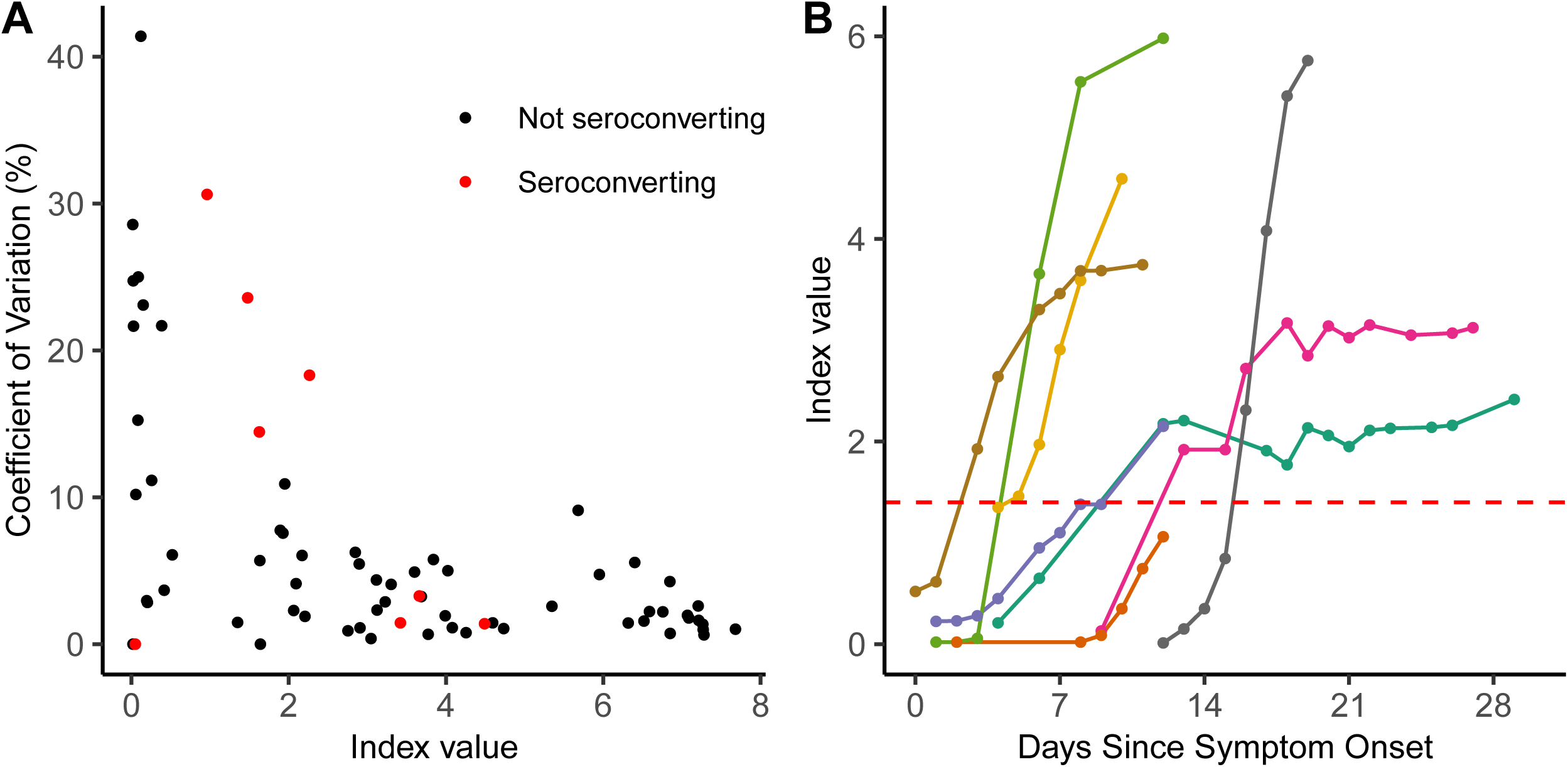
Variation among biological replicates is explained by seroconversion. A) Coefficient of variation versus index value is depicted for biological serum replicates from individuals who had more than 3 serum or plasma draws from the same calendar day. Specimens taken from individuals who were seroconverting during the repeat sampling period are shown in red. B) Index value over time since symptom onset is shown for seven individuals who seroconverted and one who failed to meet the positivity threshold during the sampling period. Each individual is depicted by a different color. The index value threshold of 1.40 for positivity is depicted in the red horizontal dashed line.

### SARS-CoV-2 Seroprevalence Survey in Boise, Idaho

We tested 4,856 individuals from Boise, Idaho sampled over one week in late April 2020 as part of the Crush the Curve initiative to determine anti-SARS-CoV-2 seroprevalence in this community. The age distribution of this cohort was: 0–19: 4.9%, 20–29: 6.2%, 30–39:17.1%, 40–49: 22.7%, 50–59: 23.5%, 60–69: 18.3%, 70–79: 6.7%, 80+: 0.5% (Table 3). The cohort had a greater representation from female individuals with 54.2% female, 41.9% male with 3.9% unknown. We detected 87 positive specimens in this cohort corresponding to a seroprevalence of 1.79%, using the manufacturer’s index value threshold of 1.40. Seroprevalence was higher among males at 2.1% than it was among females at 1.6%. Those without a reported gender had a seropositivity of 2.6%. Seroprevalence was highest among those over 80 years (4%), 60–69 year-olds (2.5%), and 20–29 year-olds (2.3%), and was lowest among those under 19 years of age (0.4%).

**Table 3.** Descriptive Epidemiology of Crush the Curve Seroprevalence Survey in Boise, Idaho

|  |  |  |
| --- | --- | --- |
|  | Total (%) | Positive (%) |
| Total | 4856 (100%) | 87 (1.8%) |
| Reported Gender |  |  |
| Female | 2631 (54.2%) | 42 (1.6%) |
| Male | 2035 (41.9%) | 40 (2.1%) |
| Unknown | 190 (3.9%) | 5 (2.6%) |
| Age (years) |  |  |
| 0-19 | 240 (4.9%) | 1 (0.4%) |
| 20-29 | 301 (6.2%) | 7 (2.3%) |
| 30-39 | 831 (17.1%) | 13 (1.6%) |
| 40-49 | 1102 (22.7%) | 18 (1.6%) |
| 50-59 | 1142 (23.5%) | 22 (1.9%) |
| 60-69 | 888 (18.3%) | 22 (2.5%) |
| 70-79 | 327 (6.7%) | 3 (0.9%) |
| 80+ | 25 (0.5%) | 1 (4%) |

## Discussion

Here we report the performance characteristics of the recently available Abbott SARS-CoV-2 IgG assay. Using the manufacturer’s recommended index value cutoff of 1.40 for determining positivity, we report an assay specificity of 99.9% from 1,020 pre-COVID-19 serum specimens and sensitivity of 100% at 17 days after symptom onset and 13 days after PCR positivity. Our results mirror that of the assay package insert, which details a 99.6% specificity from 1,000 SARS-CoV-2 specimens and 100% sensitivity by day 14 after specimens.

In our own cohort, we found increasing the threshold would have resulted in a 100% specificity and 100% sensitivity at 17 days post-symptom onset. However, the optimal threshold may depend on the intended clinical use of the test and the characteristics of the target population. Given limitations of clinical sensitivity of SARS-CoV-2 PCR testing for various sample types, IgG serology with an applied low threshold may be a useful adjunctive diagnostic for patients with negative PCR results who have been symptomatic for ≥ 7 days with a clinical presentation consistent with COVID-19 disease. In contrast, a higher threshold might be considered for PCR-negative asymptomatic patients for assessing previous undiagnosed infection. For laboratories reporting a single diagnostic result for both populations, it may be useful to report an inconclusive range corresponding to an OD ratio of roughly 0.8-1.5 with a recommendation for repeat testing to minimize false negative results associated with seroconversion. At this time, repeat serology may be preferable to a diagnostic algorithm using a secondary assay, as no specific confirmatory assay with sufficient sensitivity and specificity exists at this time.

Our serological validation was chiefly limited by use of excess serum specimens from a mostly hospitalized population known to be very recently infected with SARS-CoV-2. This convenience sample meant that PCR and serology data were not available for each day since symptom onset, requiring us to censor follow-up days accordingly (e.g. days before if the first longitudinal serological result was positive or days afterward if the last serological result was negative). The majority of patients in this study were elderly individuals – 65.6% were older than 60 years of age – many of whom also had altered mental status at time of presentation, complicating our ability to accurately ascertain symptom onset. The elderly, hospitalized population used in our sensitivity cohort could account for the delayed time to positivity seen in our cohort versus the Abbott package insert (17 vs 14 days post-symptom onset), as declining immune responses are associated with advanced age (8). It is unclear what the prevalence of antibody is in individuals with subclinical or asymptomatic infections. We were also restricted to limited descriptive epidemiological information on the serological survey conducted within the Boise, Idaho metropolitan area. The Abbott SARS-CoV-2 IgG test is also limited in that it only detects IgG antibodies directed against nucleocapsid and cannot be used for recombinant spike protein vaccine studies.

Overall, our data demonstrate excellent performance of the Abbott Architect SARS-CoV-2 IgG Assay and a high level of consistency with the package insert. Our data reinforce the limited circulation of SARS-CoV-2 in the Pacific Northwest during early 2020. We expect high-quality serological testing will be an important component of the diagnostic approach to SARS-CoV-2.

## Data Availability

Anonymized data is available upon reasonable request.

## Acknowledgements

The authors would like to thank the team behind Crush the Curve Idaho for facilitating sample collection and delivery for SARS-CoV-2 IgG testing. We would also like to thank the University of Washington Medical Center Northwest Campus clinical laboratory staff for reserving remnant serum and plasma samples from COVID-19 PCR positive patients, particularly Leanne Gilly and Michelle Manis. This work was supported by the Department of Laboratory Medicine at the University of Washington Medical Center. ALG reports personal fees from Abbott Molecular, outside of the submitted work.

